# A novel time-based surface EMG measure for quantifying hypertonia in paretic arm muscles during daily activities after hemiparetic stroke

**DOI:** 10.1101/2022.01.06.22268857

**Authors:** M. Hongchul Sohn, Jasjit Deol, Julius P.A. Dewald

## Abstract

After stroke, paretic arm muscles are constantly exposed to abnormal neural drive from the injured brain. As such, hypertonia, broadly defined as an increase in muscle tone, is prevalent especially in distal muscles, which impairs daily function or in long-term leads to a flexed resting posture in the wrist and fingers. However, there currently is no quantitative measure that can reliably track how hypertonia is expressed on daily basis. In this study, we propose a novel time-based surface electromyography (sEMG) measure that can overcome the limitations of the coarse clinical scales often measured in functionally irrelevant context and the magnitude-based sEMG measures that suffer from signal non-stationarity. We postulated that the key to robust quantification of hypertonia is to capture the “true” baseline in sEMG for each measurement session, by which we can define the relative duration of activity over a short time segment continuously tracked in a sliding window fashion. We validate that the proposed measure of sEMG active duration is robust across parameter choices (e.g., sampling rate, window length, threshold criteria), robust against typical noise sources present in paretic muscles (e.g., low signal-to-noise ratio, sporadic motor unit action potentials), and reliable across measurements (e.g., sensors, trials, and days), while providing a continuum of scale over the full magnitude range for each session. Furthermore, sEMG active duration could well characterize the clinically observed differences in hypertonia expressed across different muscles and impairment levels. The proposed measure can be used for continuous and quantitative monitoring of hypertonia during activities of daily living while at home, which will allow for the study of the practical effect of pharmacological and/or physical interventions that try to combat its presence.

## 1. INTRODUCTION

Upper extremity motor impairment after stroke is a major contributor to loss of function in daily life. Among many neurological conditions that impair control of the paretic arm, hypertonia, broadly defined as an increase in muscle tone, commonly appears in individuals with hemiplegic stroke [1, 2]. Hypertonia is clinically observed to be most prevalent in distal flexor muscles [1, 3, 4]. Cumulating evidence suggests that hypertonia present in wrist and finger muscles may be linked to the upregulation of the corticoreticulospinal system following a loss of corticobulbar projections from the lesioned hemisphere [5]. As a result, individuals with severe motor impairment after stroke often have difficulties in using their hand, for example, opening the hand or extending the wrist, which is detrimental to many activities of daily living [6]. Studies suggest that prolonged exposure to such abnormal neural drive may further impact musculoskeletal changes in the paretic arm, leading to a flexed resting posture or contractures [7, 8]. Consequently, current treatment of hypertonia largely focuses on wrist/finger and sometimes elbow flexor muscles, which include focal injection of botulinum toxin [9-11], use of wrist/finger orthosis or splints [9, 12, 13], electrical stimulation [14], stretching [15, 16], and surgical procedures such as tendon transfers [17-19].

However, there currently is no quantitative measure that can be used to constantly track how hypertonia is expressed in wrist and fingers muscles, which is crucial for understanding the true impact of hypertonia in daily function or objectively assessing the effectiveness of afore-mentioned treatments, e.g., pre and post intervention. Hypertonia in clinic is currently assessed using coarse scores such as the Modified Ashworth Scale (MAS) [20-22] and the Modified Tardieu Scale [23]. On the other hand, many studies have also attempted to assess hypertonia more quantitatively using laboratory equipment (e.g., mechatronics combined with dynamometry, sEMG) [16, 24]. However, such momentary measurements which largely relies on manual (in clinics) or instrumented (in laboratory) examination of passive resistance to joint displacement do not capture the true presence of hypertonia during daily activities. This is because daily involvement of the paretic arm is most often marked by non-use or neglect, while performing motor tasks with the non-paretic arm or the whole body [25, 26]. As such, there often is not a good correlation between these measures and patient’s ratings of abnormal tone experienced daily [11, 27, 28], and thus fail to provide useful information about the impact of hypertonia on everyday function.

Recent advances in wearable sensors, e.g., inertial measurement unit (IMU) + surface electromyography (sEMG), offer promising avenues to examine motor function of neurologically injured individuals in the real-world. As such, they have been employed in many applications in rehabilitation, e.g. to recognize motor pattern or intent for robotic prosthetics control [29-31], to detect disease symptoms or adverse events [32-34], or to provide bio-feedback during training [35]. However, there are remaining gaps in utilizing these technologies to constantly monitor an important neural signature underlying post-stroke motor impairment such as hypertonia. Most importantly, we currently lack a robust measure that can reliably quantify abnormal neural drive to paretic muscles over extended periods of time (e.g., weeks) [36, 37]. In particular, sensitivity of sEMG magnitude to noise and electrode placement makes it challenging to obtain consistent measurements across days [38-40]. For example, measurements via sEMG inherently suffers from nonstationarity of the signal, where magnitude of the signal varies with measurement condition such as electrode-skin impedance and electrode location. Further, deficits in stroke survivors to voluntarily activate their muscle raises issues in defining a reference for comparison [41-43].

In this study, we propose a novel time-based surface electromyography (sEMG) measure, Active Duration, which can overcome the limitations of the coarse clinical scales often measured in functionally irrelevant context and the challenges associated with magnitude-based sEMG measures to quantify hypertonia. We postulate that the key to robust quantification of hypertonia is to capture the “true” baseline in sEMG for each measurement session, by which we can define the length of time (i.e., duration) a muscle is active relative to this baseline using a widely accepted technique for detecting active state of a muscle from sEMG [44-46]. Such approach can be applied over a short time segment tracked in a sliding window fashion to quantify the continuous time-course of how and to what extent hypertonia is expressed, e.g., over extended period (hours) during activities of daily living. To validate the robustness of the proposed measure in real-world scenario, we first use sEMG from healthy individuals to test its robustness across parameter choices, against noise, and its reliability across measurements. Further, we demonstrate the feasibility and the utility of the proposed measure with sEMG recorded from paretic arm muscles while individuals with stroke perform various daily activities, such as, while awake, when completely relaxed, as well as during some functional activities such as ambulating in the laboratory. Results are further discussed focusing on the implications to quantifying hypertonia in paretic arm muscles during daily activities after stroke.

## 2. MATERIALS AND METHODS

### 2.1. Active duration: a time-based sEMG measure to quantify hypertonia

Our proposed time-based measure of hypertonia quantifies *Active Duration* of sEMG within a time segment, defined as the percent of time conditioned signal is above a threshold defined with signal distribution. Briefly, the method relies on robust capturing of the “true” baseline in sEMG for each measurement session using an established relaxation protocol (i.e., eliminate hyperactivity) [47, 48], from which we define the relative duration of activity over a short time segment, continuously tracked in a sliding window fashion (Fig. 1A). Specifically, four steps are involved:

**Figure 1.**
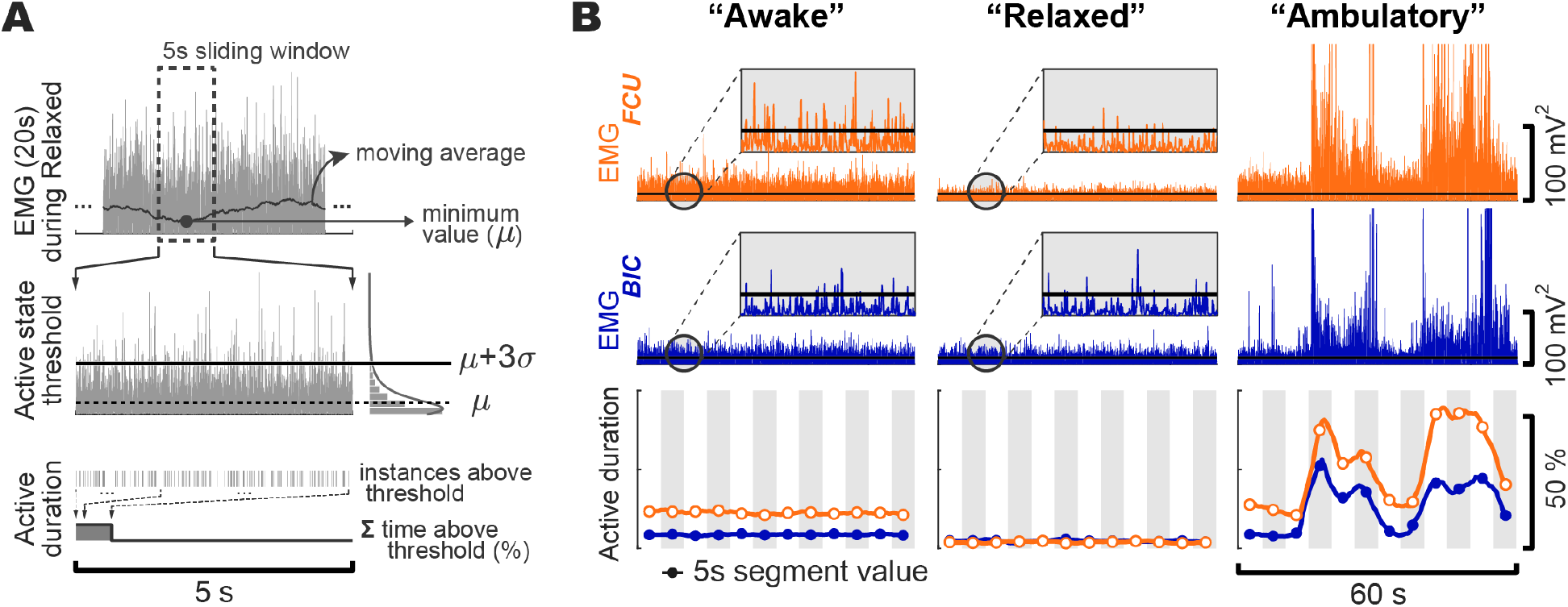
Proposed time-based sEMG measure: Active duration. (A) Conditioned sEMG is examined with a moving average window (e.g., 5 s) to find the minimum across all recordings (top), from which a threshold of active state was defined (middle). Active duration is calculated for each segment as the percent of time amplitude was above the threshold (bottom). (B) Time trace of conditioned sEMG and calculated active duration (bottom) over 60 s from a representative participant with stroke during normal *Awake*, completely *Relaxed*, and functional *Ambulatory* states.

1. Signal pre-processing and conditioning: Raw sEMG recorded from any muscle or channel can be pre-processed to first remove any noise known or identified to be present such as offset, powerline interference, or substantial motion artifact. Then, the signal is conditioned with Teager-Kaiser energy operator (TKEO) to improve signal-to-noise ratio [44-46]:

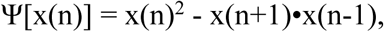

where x(n) denotes nth sample of the processed sEMG signal. Conditioned sEMG is then full-wave rectified.
2. Defining the segment-based minimum across all recordings (Fig. 1A, top): Processed and conditioned sEMG is examined with moving average window with a given length (*WL*) and overlap (in %) to find the minimum (μ) across all recordings for a given muscle. Here, *WL* and overlap are custom parameters that determine the temporal resolution of the final active duration measure (see section 2.2 below), just as any time-domain smoothing process would do, which also affects the computational demand (e.g., the smaller the *WL* and greater the overlap, the greater the demand).
3. Defining the threshold criteria (Fig. 1A, middle): For the segment where minimum mean (μ) was found above, standard deviation (σ) is calculated. The threshold criterion (*TC*) for defining active state of a muscle is defined as:

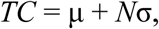

where *N* is the multiplication factor, which is a widely used method for detecting the onset in sEMG signals [44-46]. This factor *N* determines the sensitivity in detecting an active state, where ranges of numbers have been used or tested in literature [44-46], e.g., 1-3 and 3-23 for sEMG without and with TKEO conditioning, respectively. For the proposed measure active duration, *TC* determines the lower bound and the relative mapping to the signal magnitude (see section 2.2. below). While larger *TC* (e.g., μ+8σ) is often used to detect EMG onset during voluntary movements in healthy individuals [44-46], for paretic muscles where signal-to-noise ratio (SNR) is often expected to be very low [49, 50], smaller *TC* (e.g., μ+3σ) is recommended to more robustly capture the hyperactive muscle tone.
4. Active duration calculation (Fig. 1A, bottom): Once *TC* is defined from the previous step, this can be used to count, on segment basis (window length: *WL*), the number of datapoints that are above the threshold in percentage of time.

Results from segments in consideration can be then used to construct a continuous, time-course of active duration of sEMG, at given temporal resolution as determined by *WL* and overlap, where a different overlap interval can be used than in step 2), depending on the purpose. For example, one may choose to use greater overlap for more smooth and continuous visualization, whereas use results with no overlap to ensure statistical independence in analyzing time-series data [51]. An example with *WL*=5 s and *N*=3 (i.e., *TC*=μ+3σ) shows, active duration over 60 s when an individual with stroke was performing various activities of daily living (see section 2.3 for detail).

### 2.2. Validating robustness and reliability of active duration measure

To validate the sEMG active duration as a useful and reliable measure for daily monitoring or tracking of hypertonia in home-based setting, we first used experimental data from healthy individuals to systematically examine the sensitivity of sEMG active duration in detecting neural drive to muscles and quantifying the degree in relation to signal magnitude (or power). Specifically, we assessed the robustness of the proposed measure across parameter choices and against simulated noise, and its reliability across measurements conditions.

#### 2.2.1. Experimental protocol and setup

Total six healthy young adults (age 24-29 years; 3 females) participated. All participants gave informed consent prior to the experiment. All procedures and protocols were approved by the Northwestern University Institutional Review Board.

Participants completed two sessions, apart by a week on average (6.8±4.3 days), repeating the same protocol. Each day (session), participants performed series of motor tasks mimicking activities of daily living (ADL). While we ultimately seek to apply the method during any ADL, for validation purposes here, we used standardized protocols that closely mimic ADL and represent typical states of daily arm involvement and engaging sub-maximal level of muscle activation in a controlled way.

- Baseline: This task represented a completely *relaxed* state, e.g., when a participant is asleep/nearly asleep, where in the case of individuals with stroke, abnormal tone or hypertonia is expected not to be present, providing a “true” baseline. For healthy individuals, however, complete relaxation is readily achievable. During baseline, participants were seated on a standard chair with arm naturally supported on laps for 2.5 minutes.
- Timed Up and Go (TUG): This task represented the *ambulatory* state which involves a functional, whole-body movement. TUG is a clinical test designed to assess basic mobility in neurologically impaired individuals [52, 53]. The test is comprised of a sequence of functional maneuvers including rising from a chair (at the verbal cue ‘Go’), walking 3 meters (distance indicated by a line on floor to cross), walking back, and sitting down again. In the case of individuals with hemiparetic stroke, participants are allowed to use customary walking aids (e.g., cane) used in daily living.
- Timed Dumbbell (TD): This task is designed to have participants activate different groups of muscles at a controlled level, while performing a set of isotonic dumbbell-lifting (3 different weights) with their dominant (right) arm in a standardized manner, synchronized to a digital metronome with visual and audio feedback in real time. The 3 set of weights were selected for each participant, prior to the actual experiment, at levels that will engage at least perceivable effort but not inducing any fatigue and was used consistently across the two sessions for each participant. The TD tasks targeted various Degrees of Freedom (DoF), including wrist flexion/extension (WF/WE), elbow flexion/extension (EF/EE), and shoulder abduction (ABD), each repeated 7 times (see Table 1. for detailed procedure).
- Maximum Voluntary Contraction (MVC): MVCs for wrist flexion/extension (WF/WE), elbow flexion/extension (EF/EE), and shoulder abduction (ABD) were performed to capture the maximum level of voluntary muscle activation for each session. To control for consistent posture across sessions and provide support (restraint) for engaging maximal effort, participants were seated on the Biodex chair (Biodex Medical Systems, Inc., Shirley, New York, USA), with apertures fixed at similar range across participants while accommodating for each participant’s comfort (Fig. 2B), which was kept consistent across sessions. MVC trial for each DoF lasted about 5 s each, encouraged by verbal stimuli, and were repeated 3 times, with sufficient breaks in between.
- All procedures were instructed and administered by the same licensed physical therapist to ensure consistency and repeatability.

Participants repeated the sequence of Baseline, TUG, and TD three times, with consecutively increasing weights in the TD tasks. MVC trials were completed at the end of the experiment. This order was devised to minimize the influence of potential fatigue, which was also monitored through verbal communication with the participant.

**Table 1.** Target muscle list for sEMG recording

| Joints | Muscles | Abbrev. |
| --- | --- | --- |
| Wrist | *Flexor carpi radialis* | FCR |
|  | *Flexor digitorum superficialis* | FDS |
|  | *Flexor carpi ulnaris* | FCU |
|  | *Extensor carpi radialis* | ECR |
|  | *Extensor digitorum communis* | EDC |
|  | *Extensor carpi ulnaris* | ECU |
| Elbow | *Biceps brachii* | BIC |
|  | *Triceps brachii lateral head* | TRI |
| Shoulder | *Anterior deltoid* | AD |
|  | *Middle deltoid* | MD |
|  | *Posterior deltoid* | PD |

**Table 2.** Description of detailed protocols for each DoF

| DoF | TD task procedure* |
| --- | --- |
| Wrist flexion (WF) | Upper arm on side, elbow 90°, forearm supinated with palm facing superior (up), begin from fully down (extended), 7 cycles of down-up-down at 90 BPM** |
| Wrist extension (WE) | Upper arm on side, elbow 90°, forearm pronated with palm facing inferior (down), begin from fully down (flexed), 7 cycles of down-up-down at 90 BPM |
| Elbow flexion (EF) | Upper arm on side, forearm supinated with palm facing anterior, begin from elbow fully extended (0°), 7 cycles of elbow 0°-150° (fully flexed)-0° at 70 BPM |
| Elbow extension (EE) | Upper arm raised on side, forearm pronated with palm facing anterior, begin from elbow fully extended (0°), 7 cycles of elbow 0°-150° (fully flexed, behind the head)-0° at 70 BPM |
| Shoulder abduction (Abd) | Begin from arm on side, forearm in neutral with palm facing medial, keep elbow fully extended (0°) and forearm neutral, 7 cycles of shoulder 0°-90° (prior to elevation)-0° abduction at 70 BPM |

\*\* In all cases, participants were instructed to “use only as much effort needed”.
\* BPM: bit per minute.

**Figure 2.**
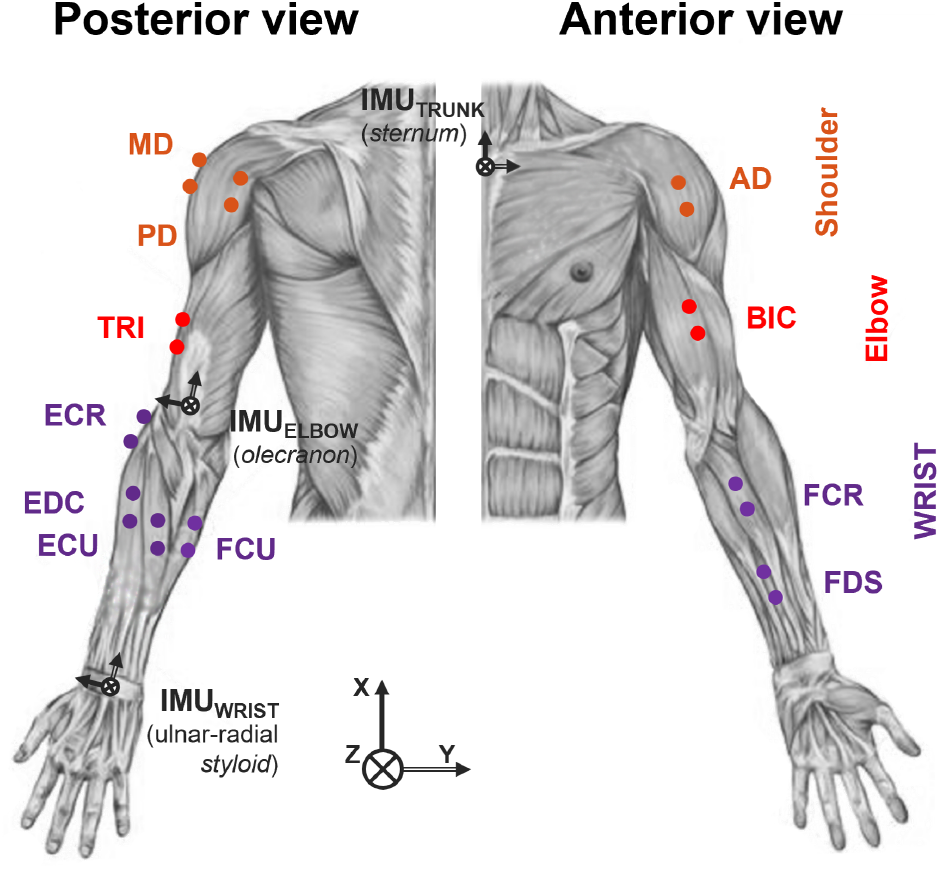
Sensor placement. Muscle activity (sEMG) was recorded from 3 shoulder (orange), 2 elbow (red), and 6 wrist/hand (purple) muscles. IMU data was recorded from two locations (trunk and wrist) to monitor gross movement and to segment sEMG data based on event detection.

During each trial in all sessions, muscle activity was recorded at 2 kHz from the given set of target muscles across the wrist/finger, elbow, and shoulder joints (see Fig. 2 and Table 1 for detailed list and abbreviations) from the dominant (right) arm using a wireless sEMG system (Delsys Trigno™, Delsys, Natick, MA). Raw sEMG signals were manually inspected to remove any artifact or noise including powerline interference in a consistent manner using an auto-regression modeling [54] and a spectral interpolation technique [55]. In addition, tri-axial acceleration and angular rate were recorded (148.148 Hz, synchronize with sEMG signal at 2:27 ratio) with inertial measurement unit (IMU) sensors attached at the trunk (sternum), elbow (proximal to olecranon fossa), and forearm (ulnar-radial styloid) (Fig. 2). IMU signals were further low-pass filtered with zero-phase Butterworth filter (4th order) at 10 Hz cut-off frequency, which were used to detect and segment sEMG data according to each task.

Between the two sessions, two different sets of sensor elements were used (assigned in random order across participants) to incorporate potential differences between elements/channels, in addition to natural inter-day variability across measurement days (see section 2.2.4).

#### 2.2.2. Robustness across parameter choices

Ranges of selected parameters involved in calculation of sEMG active duration (see section 2.1.) were tested. First, to examine the effect of window length (*WL*), which determines the temporal resolution of resulting active duration calculated, we tested *WL* of 0.5, 1, 2, and 5 s. This parameter can be also customized depending on the type of task being examine, e.g., slow/static vs. fast/dynamic, or computational demand such as memory and data storage size.

To emulate different sensor specification and re-sampling processes that may be potentially used, we tested sampling rates of 200 Hz, 400 Hz, 1kHz, and 2 kHz by decimating (with lowpass Chebyshev Type I infinite impulse response filter of order 8, to prevent aliasing during down-sampling) at ratio 1/10, 1/5, 1/2, and 1 of the original signal at 2 kHz, respectively.

To examine the effect of threshold criteria (*TC*) that defines the active state of a muscle, we tested the multiplication factor *N* (where *TC* = μ + *N*σ) of 2, 3, 5, and 10.

In all cases, when the parameter of interest was systematically varied, other parameters were kept constant, which were selected as *WL*=2 s, sampling rate = 2 kHz, and *N* = 3 as default.

Finally, we were also interested in investigating the relationship between the proposed measure and the magnitude (or power) of the signal, we examined the shape of the one-to-one mapping between the sEMG active duration and root mean square (RMS) calculated with corresponding parameter (s) on segment basis. Specifically, the initial slope (at low level) and the minimum offset of this curve was used to determine the sensitivity and consistency of sEMG active duration in detecting the onset state of a muscle, respectively. On the other hand, the range spanned and the general shape from minimum to maximum value was used to determine the extent to which and how sEMG active duration spanned the range of magnitudes (i.e., RMS).

#### 2.2.3. Robustness against simulated noises

Because our method is targeted to quantify the sEMG active duration in paretic muscles, where low SNR (i.e., due to paresis and deficit in voluntary activation [41-43]) and presence of abnormal neural drive (e.g., involuntary, sporadic firing [56]) are expected, we emulated such conditions by injecting simulated noise to the sEMG recorded from healthy individuals. In particular, we tested robustness of resulting sEMG active duration against two types of noise. First, we simulated overall increased background noise level in a muscle or a channel by injecting gaussian white noise with varying SNR 1, 2, 5, 10, and 20 dB, generated with built-in function ‘awgn.m’ in Matlab (Mathworks, Natick, MA), in which the power of the input (clean) signal is estimated using the data from representative trials of each type concatenated for each muscle/channel.

In addition, to model the sporadic motor unit firing patterns with varying statistical characteristics, we employed the model developed by Konstantin et al., [57], where parameters were modified for muscles and the sEMG data examined in this study. Briefly, the model allows for simulating synthesized sEMG signals mainly driven by the excitation profile for the motor neuronal pool, while incorporating realistic motor neuron behavior, muscle fiber innervation geometry, and tetanic force development [57]. Specifically, to test whether sEMG active duration has the sensitivity to capture the statistical characteristics of motor unit action potential, we used two types of excitation profiles (Fig. 4B, in section 3): trapezoid (15s ramp up - 15 s hold - 15s ramp down) and constant with varying firing rate (at default, 1/2, and 1/5 frequency). The synthesized EMGs were then superimposed onto the experimental sEMG data from the Baseline trials, to calculate the resulting active duration.

#### 2.2.4. Reliability across measurements

Finally, we tested the reliability of sEMG active duration measured between two different sessions, naturally affected by inter-day variability and different set of sensor elements within an individual. Reliability was assessed with an intraclass correlation coefficient (ICC, excellent when ≥0.90, good when 0.75≤ICC<0.90, moderate when 0.50≤ICC<0.75, and poor when ICC<0.50 [58]), where 3 repetitions for each type of task in each day were considered as random effect. To this end, data from each task type was segmented to consistent length and aligned in time (using IMU signals) between the two days, which encapsulated the main task in the middle. Selected lengths for Baseline, TUG, TD, and MVC trials were 65, 35, 103, and 20 s, respectively.

### 2.3. Demonstrating feasibility and utility in paretic muscles

#### 2.3.1. Experimental protocol and setup

Total three individuals with chronic hemiparetic stroke (22.0±10.5 years post stroke; age 62.7±7.5; 2 females) participated. We recruited participants who had varying levels of motor impairment, as assessed by Fugl-Meyer Assessment upper extremity portion (FMA-UE) [61], with scores apart by minimal clinically important differences [59]. All participants gave informed consent prior to the experiment. All procedures and protocols were approved by the Northwestern University Institutional Review Board.

Participants performed daily motor tasks similar to the previously described protocol (in section 2.2). However, to distinguish the baseline state when hypertonia is present or absent, we used three different states. *Relaxed* state represents when participant is asleep/nearly asleep. Following an established protocol to maximally relax paretic muscles [60], participants sat comfortably on a supported/reclined chair in a dark quiet room with relaxing musing for 15 minutes. *Awake* state represents sedentary [61, 62], yet awake condition with paretic arm at rest. Participants were seated on a standard chair with paretic arm naturally supported. *Ambulatory* state represents involvement of the paretic arm during whole-body, functional task. Participants performed the TUG test, as described before. While MVC were collected for reference, data from these trials were not included in the analysis (see below) because individuals have difficulty in voluntarily activating the muscles, especially with more severe motor impairments [41-43]. Indeed, it was found that the magnitude of muscle activity in some participants were greater when involuntary drive was present than those observed during the MVC trials. All procedures, including the clinical test (FMA-UE) were administered by licensed physical therapist.

During the experiment, sEMG and IMU data were collected as previously described, however, using a subset of muscles for sEMG, excluding the three shoulder muscles as hypertonia is less prevalent in these proximal muscles.

#### 2.3.2. Analysis

Our main focus was to test whether the proposed measure sEMG active duration can characterize the clinically observed differences in hypertonia expressed across different muscles and impairment levels well. Specifically, based on the well-known clinical observations and findings on prevalence of hypertonia [1, 3, 4], we hypothesized that active duration is greater in distal, than in proximal muscles, and increases with the severity of impairment level. To test this hypothesis, we used a linear mixed-effects model with active duration as dependent variable, and state (3 levels: *Relaxed, Awake*, and *Ambulatory*; see section 2.3.1), joint (2 levels: distal, proximal), and impairment level (i.e., FMA-UE) as fixed effects, and subject and muscle as random effects.

## 3. RESULTS AND DISCUSSIONS

### 3.1. Validation of sEMG active duration measure

For brevity and clarity, here we report and discuss only the examples from a representative participant and muscle TRI. Nevertheless, results were consistent across all muscles and all participants. All data and codes are available upon proper request to the corresponding author.

#### 3.1.1. Robustness across parameter choices

We found that the proposed measure was robust across parameter choices (Fig. 3). For example, using larger window length had smoothing effect, as would be expected for magnitude-based measure such as RMS (Fig. 3A, top left two). In relation to magnitude (Fig. 3A, right), RMS normalized to 100% at MVC, sEMG active duration sensitively (i.e., sharp initial slope) and robustly (i.e., consistent minimum offset) captured the active state of a muscle, lateral head of the triceps (TRI) in this case, while still providing a continuum of scale over the full magnitude range (i.e., range span from near-zero to 100%) in a nonlinear fashion, which asymptotically saturates at ∼25% RMS, however with a clearly distinguishable curve even beyond.

**Figure 3.**
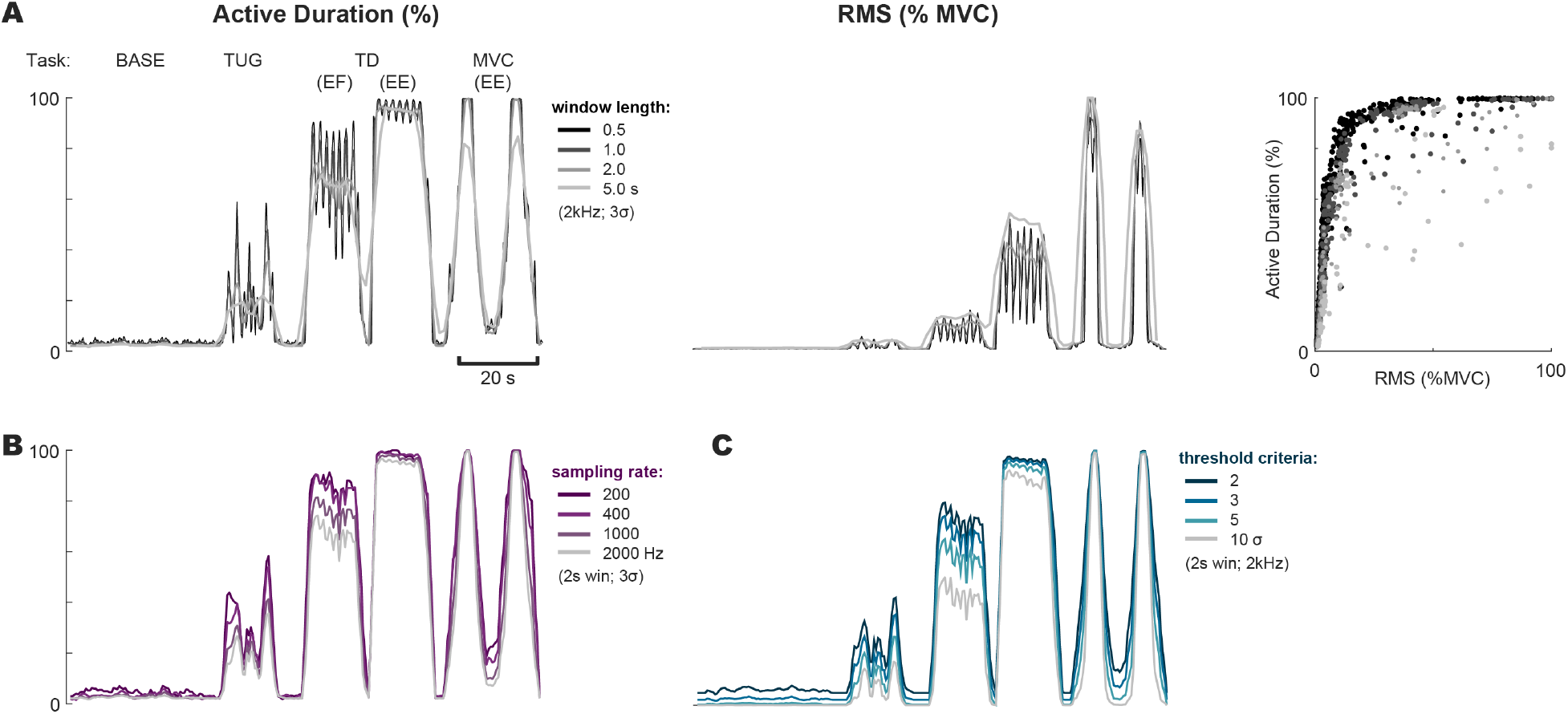
Robustness across parameter choices. (A) Resulting sEMG active duration (left) and corresponding RMS (middle) for various motor tasks, when window length is systematically varied. The relationship between active duration and RMS represented as the one-to-one mapping of active duration and RMS on segment basis (right). (B) Resulting sEMG active duration when sampling rate is systematically varied. (C) Resulting sEMG active duration when threshold criteria are systematically varied.

Testing across range of other parameters, such as sampling rate (Fig. 3B) and threshold criteria (Fig. 3B), also demonstrated that the method may allow for parameter optimization among various sensor configurations and tasks.

Robustness of the proposed measure sEMG active duration across ranges of various parameters confer reliable quantification of hypertonia across time, at both spatial and temporal resolution far exceeding the current, momentary measures in clinic- [20-22] or laboratory-based settings [16, 24]. Moreover, wider range of possible sEMG sensor configuration, e.g., low sampling rate, will allow for the use of more power-efficient sensor systems, that are more well-suited for real-world deployment, e.g., for daily use at home, outside the laboratory [32-34].

#### 3.1.2. Robustness against simulated noise sources

In addition, the proposed measure was robust against typical noise sources present in paretic muscles. When contaminated with gaussian white noise with varying SNR, the sEMG active duration could still sensitively and robustly capture active state of a muscle (Fig. 4A, left). In the case of RMS, in contrast, much of the signal was buried with low SNR (Fig. 4A, middle), which would be problematic especially when applied to muscles with substantial paresis [41-43]. Relationship to magnitude became more linear and spanned lesser range with lower SNR, however while still providing a continuum of scale that can be mapped to 0 to 100% RMS (Fig. 4A, right).

**Figure 4.**
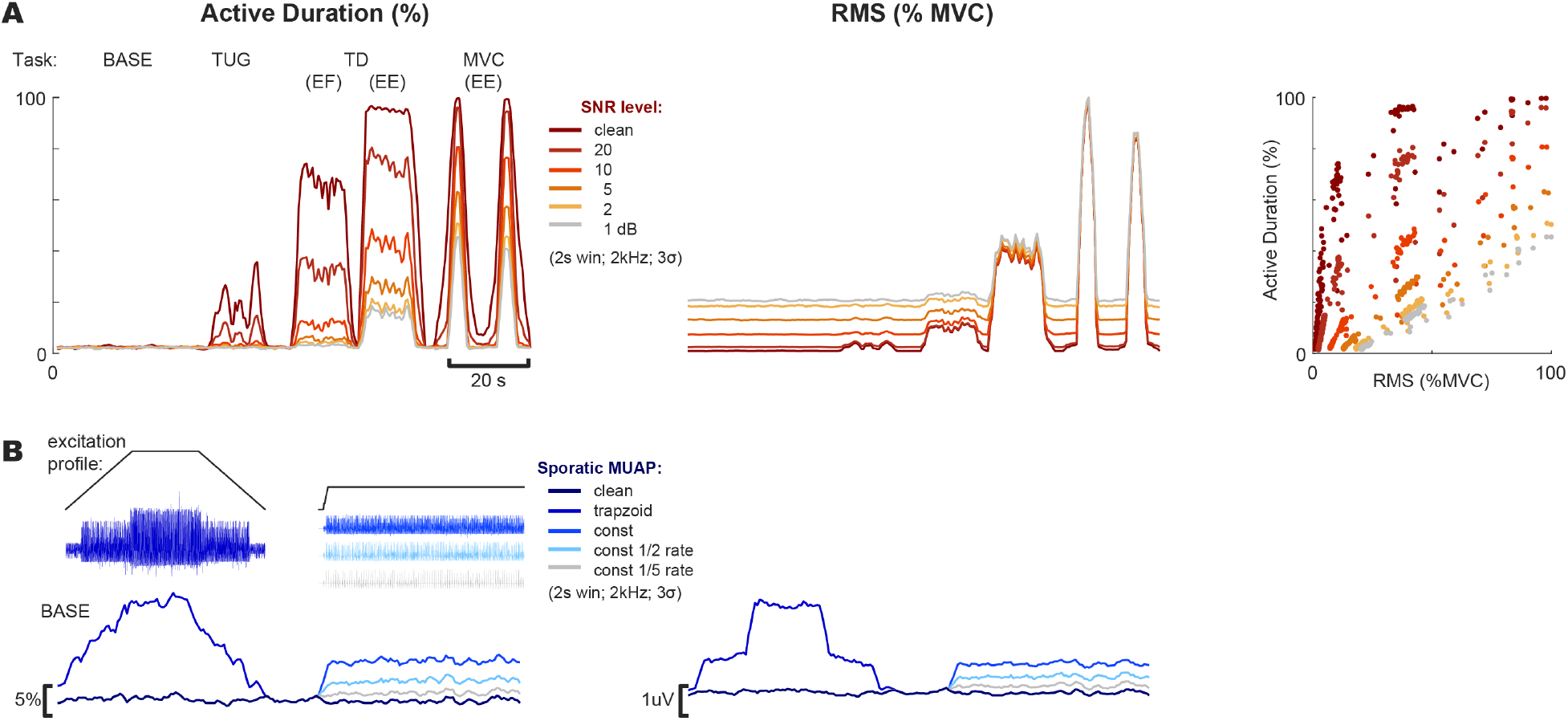
Robustness against simulated noise sources. (A) Resulting sEMG active duration (left) and corresponding RMS (middle) for various motor tasks, when background noise with varying signal-to-noise ratios (SNR) are added. The relationship between active duration and RMS (right). (B) Resulting sEMG active duration (left) and corresponding RMS (right), when simulated EMG with sporadic motor unit action potentials with varying statistical characteristics (top) are added to sEMG during baseline.

When we simulated noise that mimic sporadic motor neuron action potentials often observed in a paretic muscle, the change in temporal profile of the sEMG active duration from Baseline closely followed the excitation profile, and thus the statistical characteristics of motor unit firing or recruitment (Fig. 4B, right). The magnitude-based measure RMS, on the other hand, rather represented the amplitude of the injected noise (Fig. 4B, left), indicating that it will be subjected to ignoring the low-level manifestation of hypertonia as small offset (or noise) in background.

While pathological pattern of muscle activity has been often considered as noise or practical barriers in using magnitude-based sEMS measures [63, 64], our results suggest that sEMG active duration may be useful for examining important neuropathological characteristics of paretic muscle recruitment. Increased involuntary muscle tone (i.e., hypertonia) makes limb movement difficult and is influenced by the need/demand/intention to use the paretic/nonparetic limb or the whole body [65-67]. Evidence further suggests that the same neural mechanism, upregulation of corticoreticulospinal system due to the lost corticofugal projections [68-70], underlies the manifestation of abnormal drive to paretic muscles in various forms, resulting in the expression of the flexion synergy [71-73], spasticity [74], associated reactions [75], paresis (resulting in ‘learned non-use’ [76]) and associated functional motor deficits. Thus, provided with the exact real-world context in which such deficits are experienced and manifested, the sEMG active duration can be a useful measure to probe into the true impact of abnormal neural drive and resulting motor impairments during activities of daily living.

#### 3.1.3. Reliability across measurements

Finally, we also found that the proposed measure sEMG active duration is reliable across measurements, i.e., repetitions within a day as well as across days. For example, the time trace of 3 repetitions of the same task in the same session for each Day 1 (Fig. 5A, left) and Day 2 (Fig. 5A, right) were highly repeatable, both within days as well as across days. Inter-day reliability of sEMG active duration was also excellent, resulting in intra-class correlation coefficient over 0.99 in most muscles examined (Fig. 5B). Especially, the near-zero% value at the Baseline and near-100% value at MVC were very consistent, which was the case for all muscles and individuals. This result indicates that the measure (e.g., with default parameters selected in this study) provides highly reliable and consistent lower and upper bounds that is comparable across muscles and individuals, in stark contrast to magnitude-based sEMG measures that suffer from normalization issues [41-43].

**Figure 5.**
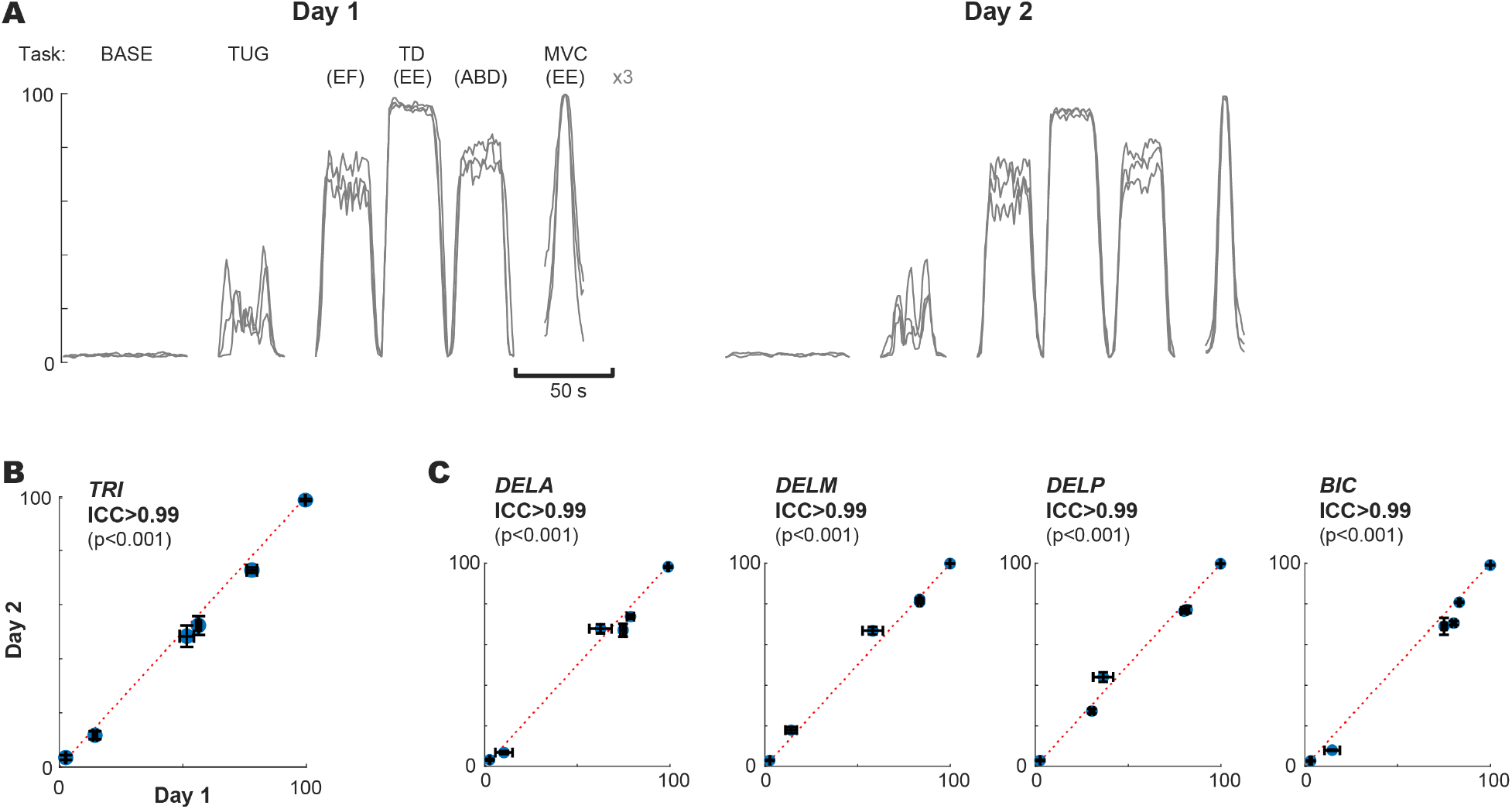
Reliability across measurements. (A) Active duration calculated for various tasks during three repetitions within a day, and across two sessions Day 1 (left) and Day 2 (right), that was apart by 3 days. Error bars indicate standard error (SE). (B) Inter-day reliability, as measured by intra-class correlation coefficient (ICC). (C) ICC for other four muscles for the same representative participant are displayed.

Furthermore, such high reliability across measurement days suggest that the sEMG active duration can be a solution to the remaining challenges in using sEMG signal to obtain robust measurements over extended period of time, e.g. across days or weeks [36, 37].

### 3.2. Feasibility and utility in paretic muscle: preliminary results

Our preliminary results (n=3) demonstrate that the proposed, time-based measure sEMG active duration can well characterize the clinically observed differences in hypertonia expressed across different muscles and impairment levels (Fig. 6). Using this measure, we found that hypertonia was more prevalent (i.e., active for longer duration) in distal wrist/finger muscles than proximal elbow muscles during *Awake* (t_254_=5.45, p<10^−6^) and *Ambulatory* states (t_282_=3.54, p<0.001). Importantly, however, there was no difference across muscles (t_254_=0.99, p=0.32) or subjects (insignificant random effect) during completely *Relaxed* state (Fig. 6A), indicating that our relaxation protocol [60] successfully quieted all muscles, and that our measure robustly quantifies the “true” baseline that is consistent/comparable across muscles and subjects, which was not the case with magnitude-based measures (e.g., RMS, normalized to MVCs). Furthermore, the amount of reduction in active duration from *Awake* to *Relaxed* state, i.e., difference between “nominal” and “true” baseline, increased as a function of impairment level (FMA-UE), more so in distal muscles (Fig. 6B; t_254_=-3.51, p<0.001), indicating that individuals with more severe impairment would experience a longer period of continuous involuntary activity, especially in the distal muscles, even during an ordinary resting state while awake.

**Figure 6.**
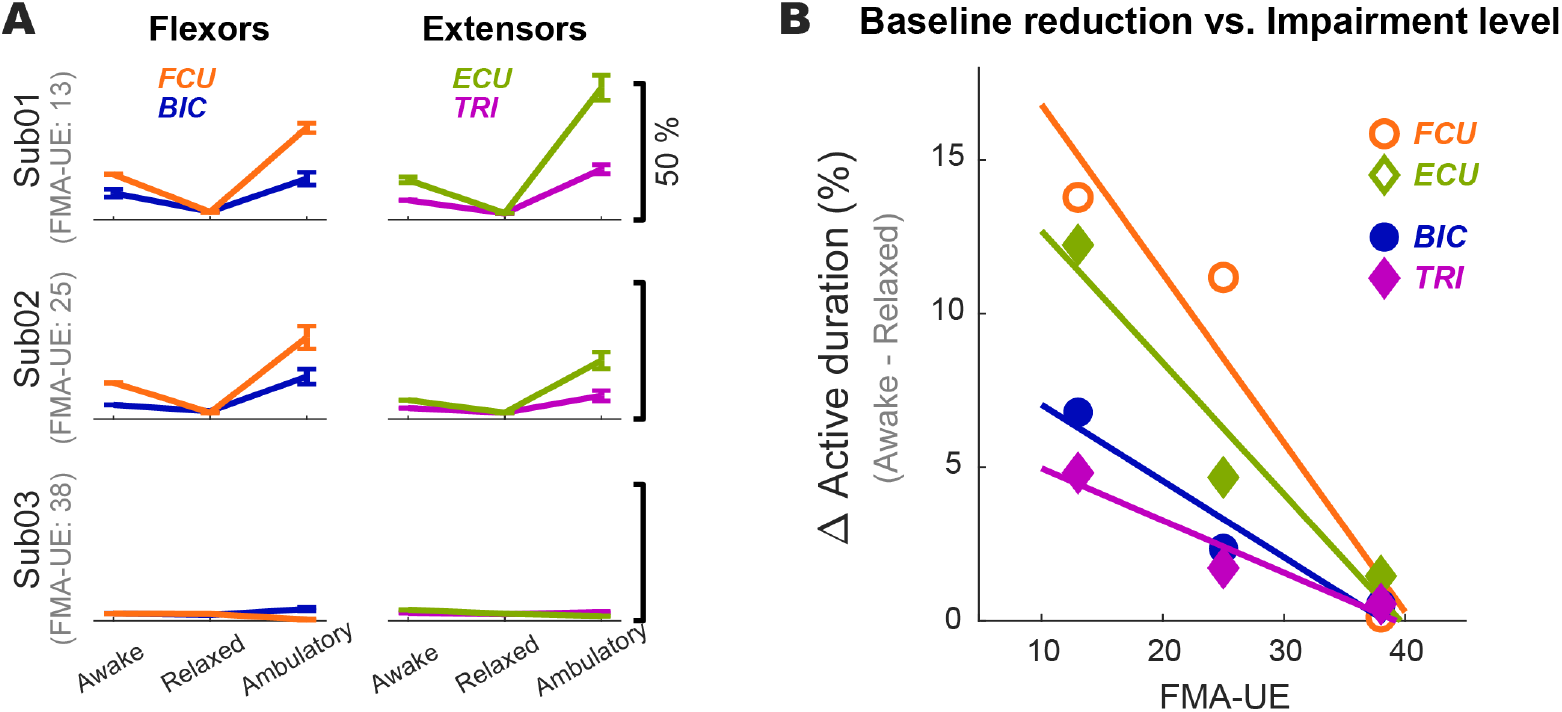
Preliminary results with individuals with stroke (n=3), with varying level of impairment (FMA-UE). (A) Comparison of sEMG active duration between distal wrist and proximal elbow muscles during *Awake, Relaxed*, and *Ambulatory* states. Each data point indicates mean±SE across 12 segments for each muscle in each state. (B) Amount of reduced time active in *Relaxed* relative to *Awake* state, as a function of impairment level (FMA-UE).

## 4. CONCLUSIONS

Here, we presented the work aimed at developing a time-based surface EMG measure that can be used to track stroke-induced hypertonia during activities of daily living. We validated that the sEMG active duration measure is robust across parameter choices (e.g., sampling rate, window length, threshold criteria), robust against typical noise sources present in paretic muscles (e.g., low signal-to-noise ratio, sporadic motor unit action potentials), and reliable across measurements (e.g., sensors, trials, and days), while providing a continuum of scale over the full magnitude range for each session. Furthermore, sEMG active duration could well characterize the clinically observed differences in hypertonia expressed across different muscles and impairment levels. Taken together, this novel time-based measure is well-suited for continuous and quantitative monitoring of hypertonia in the real-world, e.g., while at home, when integrated with commercially available wearable sensors that are easy-to-use (easy-to-”don and dof” and “wear”) [77]. Such a monitoring system will allow for the study of practical effect of pharmacological and/or physical interventions and facilitate the investigation of daily impact of hypertonia and its underlying neural mechanisms.

## Data Availability

ll data produced in the present study are available upon reasonable request to the authors.

## Supplementary Materials

N/A.

## Author Contributions

Conceptualization, M.H.S.; methodology, M.H.S. and J.D.; software, M.H.S.; validation, M.H.S. and J.P.A.D.; formal analysis, M.H.S.; investigation, M.H.S. and J.D.; resources, J.P.A.D.; data curation, M.H.S.; writing—original draft preparation, M.H.S.; writing— review and editing, M.H.S, J.D., and J.P.A.D.; visualization, M.H.S.; supervision, M.H.S. and J.P.A.D.; project administration, J.D. and J.P.A.D.; funding acquisition, J.P.A.D. All authors have read and agreed to the published version of the manuscript.

## Institutional Review Board Statement

Not applicable.

## Data Availability Statement

All data and codes used in this paper are available on proper request to the corresponding author.

## Acknowledgments

Any support given which is not covered by the author contribution or funding sections, e.g., administrative and technical support, or donations in kind.

